# Generative AI Guidelines in Korean Medical Journals: A Survey Using Human-AI Collaboration

**DOI:** 10.1101/2024.03.08.24303960

**Authors:** Sangzin Ahn

**Affiliations:** Department of Pharmacology and PharmacoGenomics Research Center, Inje University College of Medicine, Busan, Korea; Center for Personalized Precision Medicine of Tuberculosis, Inje University College of Medicine, Busan, Korea

**Keywords:** Artificial Intelligence, Scholarly Communication, Scientific Misconduct, Ethics, Research, Medical Writing

## Abstract

**Background:** Generative artificial intelligence (GAI) tools, such as large language models, have the potential to revolutionize medical research and writing, but their use also raises important ethical and practical concerns. This study examines the prevalence and content of GAI guidelines among Korean medical journals to assess the current landscape and inform future policy development.

**Methods:** Top 100 Korean medical journals by H-index were surveyed. Author guidelines were collected and screened by a human author and AI chatbot to identify GAI-related content. Key components of GAI policies were extracted and compared across journals. Journal characteristics associated with GAI guideline adoption were also analyzed.

**Results:** Only 18% of the surveyed journals had GAI guidelines, which is much lower than previously reported international journals. However, adoption rates increased over time, reaching 57.1% in the first quarter of 2024. Higher-impact journals were more likely to have GAI guidelines. All journals with GAI guidelines required authors to declare GAI use, and 94.4% prohibited AI authorship. Key policy components included emphasizing human responsibility (72.2%), discouraging AI-generated content (44.4%), and exempting basic AI tools (38.9%).

**Conclusion:** While GAI guideline adoption among Korean medical journals is lower than global trends, there is a clear increase in implementation over time. The key components of these guidelines align with international standards, but greater standardization and collaboration are needed to ensure responsible and ethical use of GAI in medical research and writing.

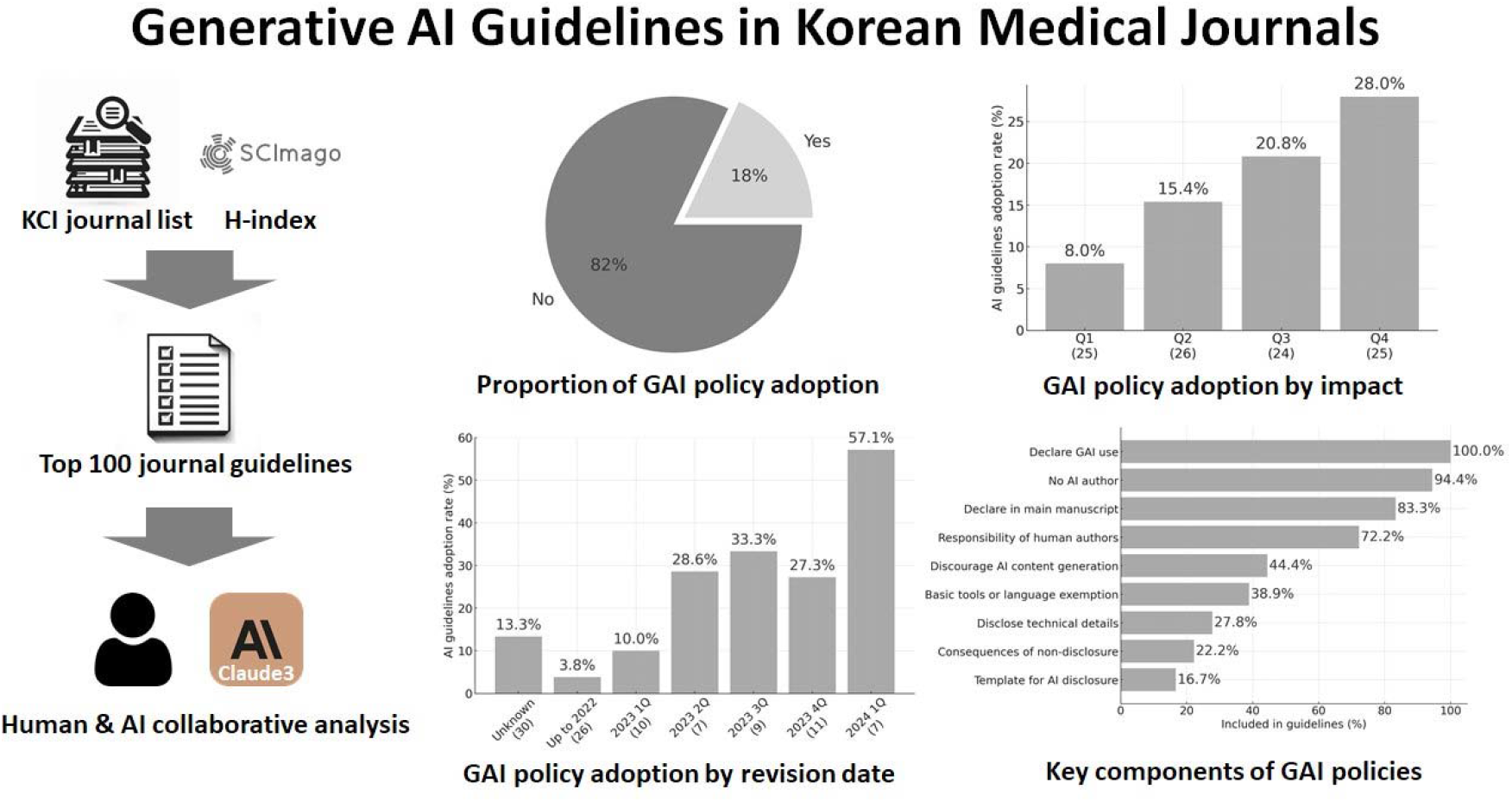

## Introduction

Generative artificial intelligence (GAI) has emerged as a groundbreaking technology with the potential to revolutionize various domains, including medical scientific publishing.^1^ GAI tools, such as ChatGPT, have gained rapid adoption and popularity due to their ability to generate human-like text based on various user prompts.^2^ In the context of medical research and academic writing, GAI offers numerous applications, such as assisting in literature reviews, data analysis, and manuscript preparation.^3,4^

For medical researchers and authors, GAI tools present several opportunities, including improving grammar and language quality, facilitating translation, generating novel research ideas, synthesizing large amounts of data, and streamlining the overall research process.^5,6^ However, the use of GAI in medical scientific writing also poses significant challenges, such as the risk of inaccuracy, bias, plagiarism, and lack of accountability.^7,8^ These concerns are particularly pressing in the medical domain, as misinformation generated by GAI could have severe consequences for patient care and public health.^9^

As the use of GAI in medical academic writing becomes more prevalent, there is a growing need for clear guidance and policies to ensure responsible and ethical practices. Medical journals and publishers have responded to this challenge with varied approaches, ranging from outright prohibition to cautious acceptance with strict disclosure requirements.^10^ The Committee on Publication Ethics (COPE) has issued a position statement on AI tools in research publications, emphasizing the importance of human authorship and responsibility while suggesting ways to disclose AI use.^11^ However, a study conducted in October 2023 that surveyed the top 100 scientific journals revealed substantial heterogeneity, with many specific guidelines not fully aligning with COPE’s recommendations.^12^

In the case of Korean medical journals, the Journal of Radiology has been actively sharing its thoughts regarding GAI and has revised its policies accordingly.^13^ Other papers have mentioned the position of GAI and its influence on medical academic writing.^14,15^ However, limited research has been conducted on the prevalence and content of GAI policies in Korean medical academic publishing. This gap in knowledge underscores the importance of examining the current landscape of GAI-related author guidelines in Korean medical journals to inform future policy development and implementation.

The present study aims to address this knowledge gap by conducting a comprehensive survey of the top 100 Korean medical journals to determine the prevalence and content of GAI-related policies in their author guidelines. Notably, this research employs a novel methodology that combines human and AI collaboration during the analysis process. By analyzing adoption rates, key policy components, and comparing findings with global trends, this study seeks to inform future policy development in Korea, fostering alignment with international standards. Ultimately, the goal is to contribute to the discourse on ethical and responsible use of GAI in medical research, ensuring its benefits are harnessed while mitigating associated risks.

## Methods

### Data Collection

The Korean Citation Index (KCI) Journal search page (https://www.kci.go.kr/kciportal/po/search/poSereSearList.kci) was used to identify Korean medical journals. Journals categorized under Medicine (의약학) and registered with the KCI (KCI 등재) were selected. A total of 538 journal entries were downloaded, resulting in 311 unique journals after removing duplicates. Journal metrics, including the H-index, were obtained from Scimago (https://www.scimagojr.com/journalrank.php). The final sample consisted of the top 100 journals by H-index, with values ranging from 3 to 104.

The official websites of the selected journals were manually searched, and the author guidelines were downloaded. Guidelines were available in PDF format for 84 journals and as webpages for 16 journals. The webpage guidelines were converted to TXT format for further analysis. The data collection process was completed within 24 hours on March 5, 2024, to ensure an accurate snapshot of the available guidelines.

### Identification of guidelines with GAI policy

The human author initially screened all 100 guidelines to identify those containing instructions related to GAI. This process yielded 18 guidelines with GAI-related content. The guideline files (PDF or TXT) were uploaded to an AI chatbot (Claude 3 Opus) to verify the presence of GAI-related content. The results of the human and AI identification processes were compared, and no discrepancies were found.

The last revision date for each guideline was also recorded during the screening process. If the revision date was not available, it was marked as “Unknown.” A total of 70 guidelines had a revision date, while 30 did not. The analysis process is depicted in Fig. 1.

**Fig. 1.**
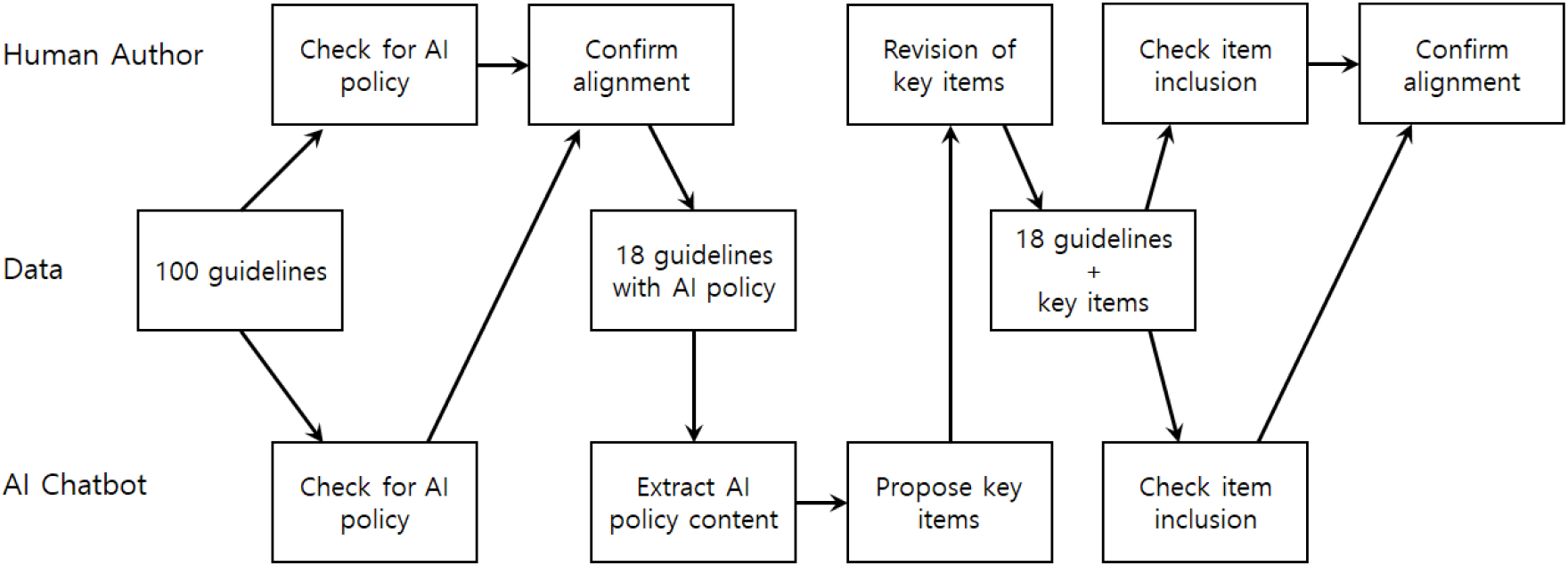
Evaluation workflow for AI policy in author guidelines. Initially, both a human author and an AI chatbot verify the existence of AI policies within 100 guidelines. From the 18 guidelines identified as having AI policies, the AI chatbot extracts AI-related content and proposes key items for evaluation. After revision of the key items by the human author, the inclusion of these items in the 18 guidelines is checked by both the human and AI. Final confirmation of item inclusion is performed by the human author.

### Content analysis of GAI-related items in guidelines

The AI chatbot was used to identify and extract GAI-related content from each of the 18 guidelines that have adopted GAI policy. All extracted content was again uploaded to the AI chatbot, which then suggested key items for comparison across journals. The human author examined the AI-suggested key items and revised them into 11 key items for comparison across journals.

The human author manually assessed the presence of the 11 key items in each of the 18 GAI-related guidelines. The AI chatbot was also provided with each of the 18 guidelines and asked to check for the presence of the 11 key items, answering “YES,” “NO,” or “UNSURE” for each item. Any discrepancies between the human author and AI chatbot assessments, as well as items marked “UNSURE” by the chatbot, were reexamined by the human author. A total of 5 discrepancies were observed: 2 in “Emphasize human responsibility,” 2 in “Prohibition of AI usage other than language improvement,” and 1 in “Declare in manuscript.” All other items tagged “UNSURE” were determined to be “NO” by the human author.

### Analysis of journal characteristics and GAI guideline adoption

Journals were divided into quartiles based on their H-index values. The percentage of journals with GAI guidelines in each quartile was calculated. Journals were also grouped according to the last revision date of their guidelines: Unknown, Up to 2022, 2023 1Q, 2023 2Q, 2023 3Q, 2023 4Q, and 2024 1Q. The percentage of journals with GAI guidelines in each group was calculated.

### AI chatbot usage

In this research, Claude 3 Opus, the most intelligent model developed by Anthropic (CA, USA) as of March 2024, was employed. This model, part of the Claude 3 series which also includes Sonnet and Haiku, is distinguished by its ability to navigate open-ended prompts and sight-unseen scenarios with remarkable fluency and human-like understanding.^16^ The AI chatbot was used to identify and extract GAI-related content in guidelines, suggest key items for analysis, and check for the inclusion of each item in the author guidelines. Additionally, it was utilized during manuscript writing to suggest ideas and improve the linguistic quality.

### Statistical analysis and visualization

Percentages were calculated to summarize the prevalence and characteristics of GAI guidelines among the selected Korean medical journals. The adoption of GAI policies was calculated overall, by H-index quartile, and by revision date. All calculations and visualizations were initially performed using Data Analyst by ChatGPT (OpenAI, CA, USA), an AI-powered data analysis tool. To ensure the accuracy and reliability of the results, the human author subsequently confirmed the findings using the Matplotlib package (version 3.7.1) in the Python language (version 3.10.12) within a Google Colaboratory environment (Google Research, CA, USA).

## Results

### Prevalence and Trends of AI Guideline Adoption in Korean Medical Journals

The survey of 100 Korean medical journals revealed that only 18% had guidelines addressing the use of generative artificial intelligence (GAI) tools in the research and writing process (Fig. 2A). The majority of journals (82%) lacked any formal policies or guidance on this topic.

**Fig. 2.**
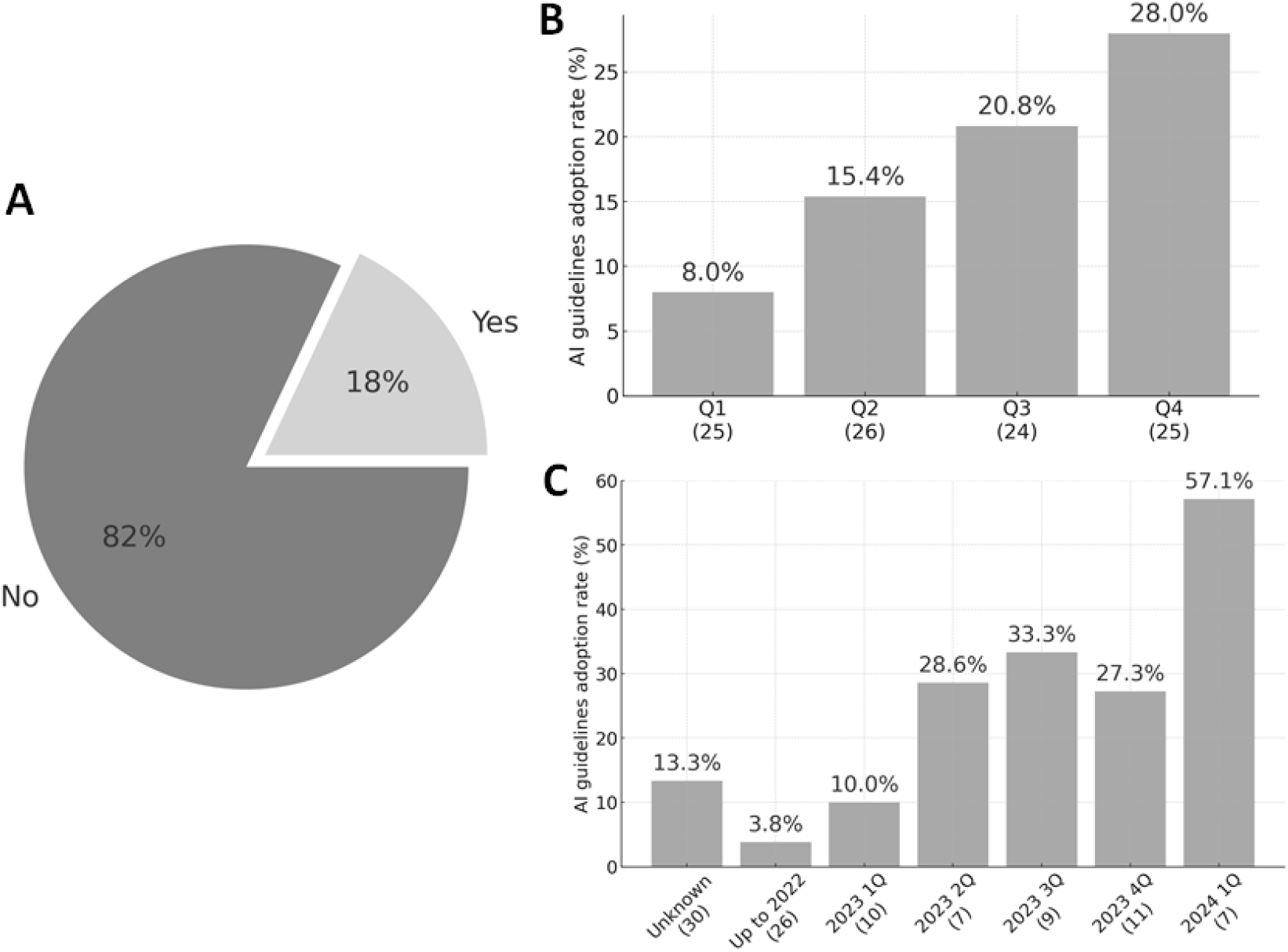
Prevalence and trends of AI guidelines adoption in Korean medical journals. (**A**) Proportion of journals with (18%) and without (82%) AI guidelines. (**B**) Proportion of journals with AI guidelines by H-index quartile. Results show higher adoption rates in journals with higher impact (Q1: 8.0%, Q2: 15.4%, Q3: 20.8%, Q4: 28.0%). (**C**) Proportion of journals with AI guidelines by revision date. Results show increasing adoption of AI guidelines over time, with 57.1% of journals implementing such policies in the first quarter of 2024, compared to only 10.0% in the first quarter of 2023.

Further analysis of the journals’ H-index quartiles showed a trend of higher GAI guideline adoption rates among journals with greater impact (Fig. 2B). The adoption rate was lowest in the first quartile (Q1) at 8.0%, increasing to 15.4% in Q2, 20.8% in Q3, and reaching 28.0% in the highest impact quartile (Q4).

Examining the temporal trends in GAI guideline adoption, a clear increase was observed over time (Fig. 2C). Among journals with unknown revision dates, 13.3% had GAI guidelines. This proportion was only 3.9% for journals with guidelines last revised before 2023. However, the adoption rate increased to 10.0% in the first quarter of 2023, 28.6% in the second quarter, 33.3% in the third quarter, and 27.3% in the fourth quarter. Notably, 57.1% of journals that revised their guidelines in the first quarter of 2024 had incorporated GAI policies.

### Key Components of AI Policies in Korean Medical Journal Guidelines

Among the 18 journals with GAI-related guidelines, all required authors to declare the use of GAI tools in some form (Fig. 3). Most journals (94.4%) explicitly stated that GAI tools could not be listed as authors due to the lack of AI’s ability to take responsibility. The majority of the journals (72.2%) also emphasized the responsibility of human authors for the scientific integrity and content of the manuscript.

**Fig. 3.**
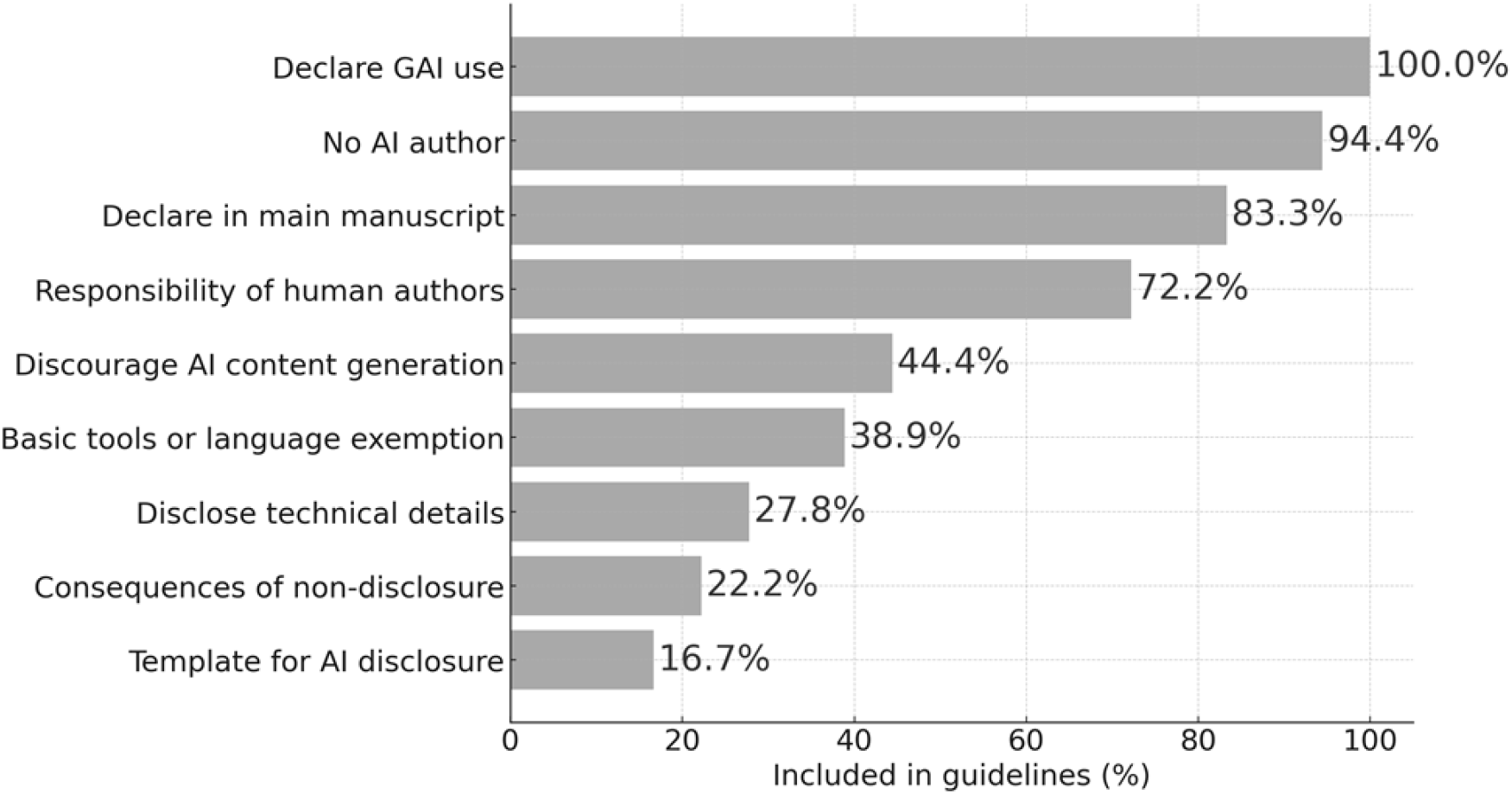
Key components of AI policies in Korean medical journal guidelines. All journals require authors to declare the use of GAI in their manuscripts, and nearly all (94.4%) prohibit listing AI as an author. A majority of the journals (83.3%) require authors to declare AI use in the main manuscript, and 72.2% emphasize the responsibility of human authors for the paper’s content. Less than half (44.4%) discourage the use of AI for content generation, while 38.9% exempt the use of basic tools or AI for language improvement from the declaration requirement. Fewer journals require disclosure of technical details (27.8%), specify consequences for non-disclosure (22.2%), or provide a template for AI disclosure statements (16.7%).

Regarding the location of the GAI declaration, 15 journals (83.3%) required it to be made in the main manuscript, with variations in the specific section: 4 in the materials and methods section, 3 in the acknowledgements or materials and methods section, 3 in a new dedicated section, 1 in the acknowledgements section, and 4 did not specify the section name. Eight journals (44.4%) discouraged the use of GAI tools for content generation, with 3 stating that AI tools should only be used to improve language quality and 5 prohibiting the use of AI tools to generate text, images, or figures. Seven journals (38.9%) mentioned exemptions from the declaration requirement for the use of basic tools or GAI solely for enhancing linguistic quality, with 5 specifying that “traditional” basic tools for grammar, spelling, and references do not need declaration, and 2 explaining that “modern” tools can be used without declaration for language improvement.

Less common policy components included the requirement to disclose technical details of the GAI models used, such as name, version, and manufacturer (27.8%), statements on the consequences of non-disclosure, such as rejection or retraction (22.2%), and the provision of templates for GAI usage disclosure statements (16.7%).

## Discussion

This study investigated the prevalence and content of generative artificial intelligence (GAI) guidelines among the top 100 Korean medical journals. The findings reveal that only 18% of these journals have implemented policies addressing the use of GAI tools in research and writing. Although this adoption rate is lower than the global prevalence of 87% among top scientific journals, as reported by Ganjavi et al., it is crucial to recognize the increasing trend of GAI guideline adoption in Korean journals over time, particularly in the first quarter of 2024.^12^ These results suggest that Korean medical journals may be in the earlier stages of adopting GAI policies compared to global leaders in the field.

Comparing the key components of GAI guidelines in Korean journals to the common themes identified in global studies by Ganjavi et al. and Inam et al. reveals several similarities.^12,17^ The universal prohibition on listing AI as an author and the requirement for authors to disclose AI use are consistent across Korean and international journals. Additionally, the shared emphasis on human author responsibility for manuscript content aligns with the recommendations set forth by the International Committee of Medical Journal Editors (ICMJE) and the COPE.^11,18^ These core principles underscore the importance of maintaining human oversight and accountability in the use of GAI tools in medical research and writing.

Despite the similarities in key principles, there are notable differences in the specificity and structure of GAI guidelines between Korean and global journals. Korean journals tend to have less detailed requirements regarding what information should be disclosed when using GAI tools, whereas international journals often provide more comprehensive guidance. Furthermore, there is significant variability among Korean journals in terms of where and how GAI use should be disclosed within the manuscript. The rates of requiring technical details, providing disclosure templates, and specifying consequences for non-compliance also vary. These findings highlight the need for greater standardization of guideline content and structure among Korean medical journals, a challenge that is also present in the global context.

Although not noted in the results, some author guidelines (3 of 18) mentioned that AI tools must not be used for the peer review process. This aligns with the findings of Inam et al., which highlighted the strict prohibition of AI use in peer review among the top 25 Cardiology and Cardiovascular Medicine journals. The prohibition of AI in peer review is due to concerns regarding confidentiality and the need for expert human insight in evaluating scientific work. However, it is important to note that the guidelines for reviewers were not assessed in this study, and the policies regarding the use of AI tools during the review process may warrant further investigation in future research.

The results of this study underscore the need for Korean medical journals to continue expanding GAI policy adoption, particularly among lower-impact journals. To address the heterogeneity in guideline content and structure, it is recommended that Korean journals collaborate with global initiatives to develop cohesive, cross-disciplinary standards for GAI use in medical research and writing. Additionally, author education and editorial oversight are crucial to ensure compliance with these guidelines. As GAI capabilities continue to evolve rapidly, regular updates to the guidelines will be necessary to keep pace with technological advancements and emerging ethical concerns.

It is important to acknowledge the limitations of this study, such as the focus on a limited number of high-impact Korean medical journals and the lack of assessment of reviewer guidelines. Future research should examine GAI policy implementation, author compliance, the impact of these policies on the peer review process, and the use of AI tools in peer review. As GAI tools become more sophisticated and widely adopted, it will be essential to investigate their influence on the quality, integrity, and efficiency of medical research and publishing.^19,20^

In conclusion, this study provides valuable insights into the current state of GAI guidelines among Korean medical journals. While the adoption rate is lower than global trends, there is a clear increase in the implementation of GAI policies over time. The key components of these guidelines align with international standards, but greater standardization and collaboration are needed to ensure responsible and ethical use of GAI in medical research and writing. As GAI tools continue to advance and transform the landscape of medical research, it is crucial to establish clear, comprehensive, and cohesive policies that promote transparency, integrity, and the pursuit of scientific knowledge.

## Data Availability

All data produced in the present study are available upon reasonable request to the author

## Supplemental information

Prompts used in this study.

### 1. Checking presence of GAI related content in the guideline

Prompt:

The attached file is the author guideline for a scientific journal. Extract the contents related to instructions to authors on use of generative artificial intelligence or language models. Write a summary at the end.

Example AI response:

The author guidelines include specific instructions regarding the use of generative artificial intelligence (AI) in the “Ethics and Authorship in the Use of Generative Artificial Intelligence (AI)” section:

- Assigning authorship to AI is prohibited.
- Authors who use generative AI tools are solely responsible for all content produced and submitted.
- The journal discourages using generative AI tools to create any content for scientific manuscripts. If such tools are used, authors must transparently report their use.
- Using AI tools to enhance the linguistic quality of a submission is considered acceptable and does not require specific disclosure.
- When generative AI is the focus of a study, its use should be explicitly detailed in the Materials and Methods section.
- Reviewers are forbidden from using AI to generate review comments.

In summary, the journal prohibits granting authorship to AI, discourages the use of generative AI for creating manuscript content, and requires transparent reporting if such tools are used. Using AI for language enhancement is allowed without disclosure. AI should be described in the methods if it is the study focus. Reviewers cannot use AI to write comments.

### 2. Extracting GAI related content from each guideline

Prompt:

The attached file is the author guidelines for a scientific journal. Extract the contents related to instructions to authors on use of generative artificial intelligence or language models. Answer straight to the point and include all information. Organize into bullet points. Refrain from adding introductory statements in the beginning and just answer in bullet points.

Example AI response:

- If artificial intelligence (AI)-assisted technologies (such as large language models, chatbots, or image creators) were used during preparation of the manuscript, authors should disclose how they used AI in both the cover letter and the submitted work.
- Authors are responsible for scientific integrity of all contents that AI generated.
- Authors should not list AI-assisted technologies as an author or co-author, nor cite AI as an author.
- All submissions or publications that do not adequately disclose AI assistance will be rejected from review or retracted.

### 3. Proposing key items for further comparative analysis

Prompt:

The attached file includes contents from 18 journal submission guidelines that are related to use of generative artificial intelligence or language models. Break down into small items that can be used to compare between journals. Write the items in bullet points.

AI response:

Here are the key items extracted from the 18 journal submission guidelines, broken down into bullet points for comparison:

- Authorship:
  - AI/LLMs cannot be listed as authors or co-authors
  - AI/LLMs cannot be cited as authors
  - Authorship implies responsibilities that can only be attributed to humans
  - Authors are responsible for the scientific integrity of AI-generated content
- Disclosure:
  - Use of AI/LLMs must be disclosed in the cover letter and manuscript
  - Specific tools, models, versions, and manufacturers should be detailed
  - Disclosure should explain how AI was used and in what capacity
  - Required location for disclosure varies (e.g., Acknowledgments, Materials and Methods)
  - Basic tools for grammar, spelling, and references may not require disclosure
- Acceptable use:
  - AI/LLMs can be used to improve readability and language
  - Use should involve human oversight, control, review, and editing
  - Authors should carefully review AI output for accuracy, bias, and completeness
- Unacceptable use:
  - AI/LLMs should not be used to write manuscripts (with some exceptions)
  - Using AI for creating/altering figures, images, and artwork is generally discouraged
  - If AI is used for figures/images, clear descriptions must accompany the submission
- Plagiarism and attribution:
  - Authors must ensure no plagiarism in AI-generated text and images
  - Appropriate attribution and citations are the responsibility of human authors
- Consequences of non-disclosure:
  - Submissions without proper AI disclosure may be rejected or retracted
  - Previously published papers may be retracted if AI use is later discovered
- Reviewer guidelines:
  - Reviewers should not use AI when reviewing manuscripts
  - Sharing manuscripts with external AI tools during review can break confidentiality

### 4. Check for inclusion of items in each guideline

Prompt:

Read the attached author guidelines and answer ‘YES’ or ‘NO’ or ‘UNSURE’ if the author guidelines have contents related to instructions for authors on the use of generative artificial intelligence, such as language models or generative AI tools. Answer number 11 with the name of the section. Before giving the answer for each item, provide a verbatim quote from the author guidelines that is relevant to the item, if applicable. If there is no relevant content for an item, write ‘No related content’.

Items:

1. No authorship is allowed for generative artificial intelligence
2. Emphasis on responsibility for human authors on AI generated content
3. Prohibits AI usage other than language improvement
4. Use of AI must be disclosed in some form
5. Specific technical details of the AI model used should be declared
6. AI declaration template provided
7. Discourage AI usage for content generation, such as text, figure, images
8. Non-disclosure consequences are mentioned
9. Basic tools for grammar, spelling, references do not require disclosure
10. Use of AI must be declared in the cover letter
11. Use of AI must be declared in the main manuscript (If YES, specify which section: ex: acknowledgement, materials and methods, specific section, not specified. If NO, write ‘NO’).
12. Provide any other relevant information from the author guidelines related to the use of AI or language models that doesn’t fit into the items above. If there is no additional relevant information, write ‘NONE’. Instructions:

In the answer, DO NOT repeat the items. No introduction statements in the beginning. Just provide the relevant quote and then the answer. Start writing with ‘1.’.

Example AI response:

1. “Large Language Models (LLMs), such as ChatGPT, do not currently satisfy our authorship criteria. Notably an attribution of authorship carries with it accountability for the work, which cannot be effectively applied to LLMs.” YES
2. No related content. UNSURE
3. No related content. NO
4. “Use of an LLM should be properly documented in the Methods section (and if a Methods section is not available, in a suitable alternative part) of the manuscript.” YES
5. No related content. NO
6. No related content. NO
7. No related content. NO
8. No related content. NO
9. No related content. UNSURE
10. No related content. NO
11. “Use of an LLM should be properly documented in the Methods section (and if a Methods section is not available, in a suitable alternative part) of the manuscript.” YES (Methods section or a suitable alternative part if Methods section is not available)
12. NONE

